# Human Intuition vs. Computational Precision: Neurologists, Feature-based Models, and Deep Learning for Stroke Prognosis

**DOI:** 10.64898/2026.06.12.26355559

**Authors:** Lisa Herzog, Nelly Blindenbacher, Christoph Globas, Marcellina Isabelle Haeberlin, Philipp Baumgartner, Francesco Capecchi, Corinne Inauen, Beate Sick, Charles Majoie, Wim van Zwam, Susanne Wegener

## Abstract

**Background:** Prognostication in large vessel occlusion (LVO) stroke remains challenging. Although several prognostic models exist, their comparison to clinician performance, human-model interaction, and specific sources of human bias remain poorly understood.

**Methods:** Using pre-treatment clinical and CT data from the MR CLEAN trial (n=500), six neurologists predicted three-month modified Rankin Scale (mRS) scores for 40 patients, both unaided and assisted by a validated feature-based model (MR PREDICTS). Human performance was benchmarked against MR PREDICTS and a multimodal, interpretable deep learning (DL) approach using raw imaging data. We explicitly assessed neurologists’ ability to estimate model-required imaging features and identified systematic human biases. Models were additionally validated in a larger MR CLEAN trial cohort (n=404).

**Results:** For predicting the full mRS distribution, standalone models achieved good ordinal agreement (MR PREDICTS quadratic weighted kappa (QWK) 0.51 [0.24 to 0.70]; DL model 0.49 [0.25 to 0.67]), significantly outperforming unaided neurologists (QWK 0.27 [0.10, 0.42]). Neurologists showed systematic overoptimism, predicting lower mRS scores than observed. Furthermore, there was poor accuracy in extracting imaging features. Raters’ ASPECTS predictions deviated by 3.4 points from the confirmed scores, and collateral score accuracy was 44.6%. However, for predicting binary mRS (0-2 vs. 3-6), accuracy was comparable between unaided neurologists (64.17% [55.42% to 72.92%]) and models (MR PREDICTS 67.50% [52.50% to 82.50%]; DL model 63.16% [47.37% to 78.95%]). Model-assistance modestly improved and harmonized neurologists’ predictions (QWK 0.41 [0.22 to 0.55]; binary accuracy 68.75% [58.33% to 78.34%]. Model performance remained robust in the larger cohort.

**Conclusions:** Multimodal prognostic models outperform clinicians in predicting the full range of mRS outcomes, while human error in imaging assessment and systematic optimism bias are primary drivers of prognostic inaccuracy. End-to-end DL models eliminate human-input variability and hold strong potential as an automated second opinion to support prognostication and decision-making in acute LVO stroke.

## Introduction

While endovascular treatment (EVT) of patients with large vessel occlusion (LVO) stroke has significantly improved outcomes in multiple randomized controlled trials [3, 4, 5, 6, 7], patient trajectories after EVT remain highly heterogeneous. Only about half of treated patients achieve functional independence at three months [5] while individualized prognosis and potential treatment benefit are difficult to predict [8].

In clinical practice, neurologists analyze a vast amount of clinical and imaging data to estimate potential functional outcome, with and without treatment, for acute decision-making and communication with patients and their relatives. However, even in controlled experimental settings, experienced clinicians have been shown to struggle with accurately predicting functional outcome at discharge [9], three months [10, 11] and six months [12, 13] when being provided with pre-treatment clinical data. Furthermore, while imaging is fundamental to acute stroke care, the addition of Magnetic Resonance Imaging (MRI) data has not been found to improve human predictive performance but instead increased interrater variability [11].

Several prognostic models for functional outcome prediction, typically measured by the three-months modified Rankin Scale (mRS), have been proposed to support clinicians. Traditional regression-based approaches such as THRIVE [14] and DRAGON [15] or MR PREDICTS [16] combine clinical variables with selected imaging features, and were shown to outperform medical experts in controlled prediction experiments [10, 9, 13, 17], particularly when imaging data was added [11]. However, their real-world clinical utility depends on the accurate and timely assessment of their inputs, including imaging-derived features such as ASPECTS. In acute settings, such manual assessments are often hindered by significant interrater variability, human error, and time constraints which may introduce inconsistency into the prognostic chain.

Recent multimodal machine learning, particularly deep learning (DL), models offer an alternative by directly integrating clinical variables with raw imaging data, eliminating the need for manual feature extraction [18, 19, 20, 21, 22]. These end-to-end approaches have demonstrated promising performance compared with both feature-based models [23] and clinicians [11] when imaging data was incorporated into prediction. However, their clinical value remains uncertain, particularly regarding their performance relative to clinicians when being provided with clinical and Computed Tomography (CT) data. Importantly, how model-derived predictions influence clinician judgment, and whether clinicians can reliably provide the manual inputs that traditional models require, remains underexplored in LVO stroke.

In this study, we systematically evaluated neurologists’ ability to predict three-month functional outcome in patients with LVO stroke using pre-treatment clinical and multimodal CT imaging data from the MR CLEAN trial. In 40 cases, we compared neurologists unaided predictions to those with MR PREDICTS assistance, while explicitly assessing human accuracy in estimating model-required imaging features. Human judgment was benchmarked against MR PREDICTS alone and an interpretable, multimodal DL model [8]. To contextualize model performance under ideal input conditions and assess the robustness of the models in a larger patient cohort, both MR PREDICTS and the DL model were additionally evaluated in complete cases of the MR CLEAN trial (n=404). By analyzing prediction accuracy, variability, and human-model interaction, this study aimed to identify the specific drivers of the gap between human intuition and computational precision to inform the future development of robust and clinically useful prognostic tools for acute care (Figure 1).

**Figure 1:**
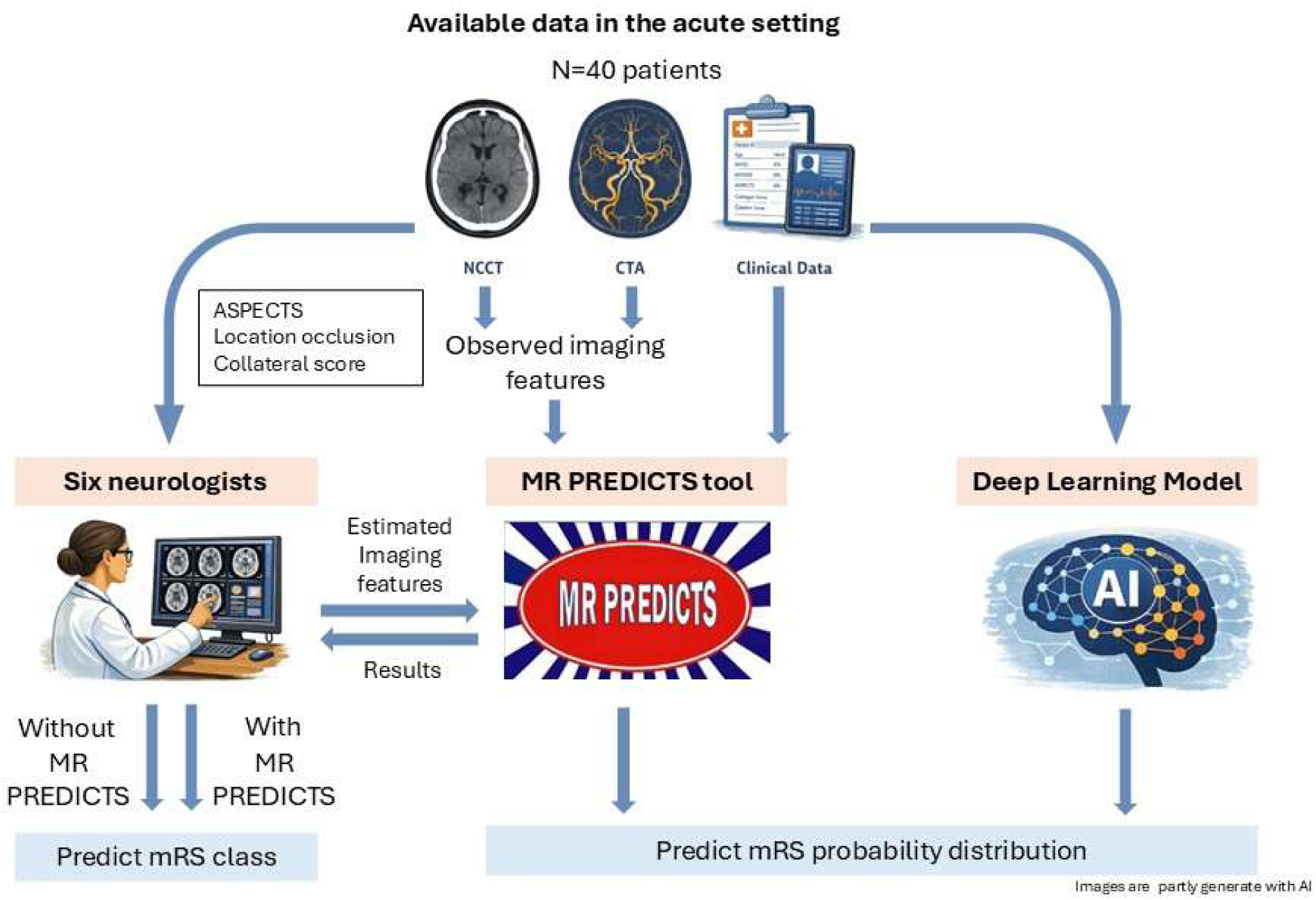
Illustration of the study. Based on non-contrast CT (NCCT), CT angiography (CTA) and clinical data, six neurologists predicted three-months mRS class (i) without prognostic support (ii) with access to MR PREDICTS. For the MR PREDICTS, neurologists were required to estimate imaging features including ASPECTS, occlusion location and collateral score. Performance of neurologists was compared to MR PREDICTS alone and to a DL model which integrated raw CTA imaging alongside clinical features.

## Methods

This study is reported in accordance with the TRIPOD guidelines [24] and relevant CONSORT-AI reporting principles [25].

### Study design and data source

This prognostic study used patient data from the MR CLEAN randomized controlled trial [4], enrolling 500 patients with imaging-confirmed LVO stroke who were randomly allocated to EVT (n=233) or no EVT (n=267). No advanced neurovascular imaging selection criteria were applied, resulting in a heterogeneous cohort receiving treatment within six hours after symptom onset. Alongside established clinical features, an imaging committee determined imaging features including ASPECTS on non-contrast CT, location occlusion on CTA, MRA or DSA and collateral score.

### Standard Protocol Approvals and Patient Consents

The trial was approved by a central medical ethics committee and the research boards of the participating centers. Before randomization, all participants (or their legal representatives) have provided written informed consent [4]. The secondary usage of the data for this study complies with data protection regulations and applicable ethical standards.

### Evaluated models

We evaluated two distinct prognostic approaches to compare the impact of manual feature extraction against automated end-to-end analysis.

The first, MR PREDICTS, is a validated, publicly available decision support tool that represents the current standard for feature-based modelling in LVO stroke (https://www.contrast-consortium.nl/mr-predicts/). This ordinal regression-based model estimates the three-month mRS probability distribution with and without EVT, as well as treatment benefit, using manually derived CT imaging features (occlusion location, ASPECTS, and collateral score) alongside routine pre-treatment clinical variables. The model was originally developed using MR CLEAN trial data [16] and updated with MR CLEAN registry data [26], making the MR CLEAN trial cohort a favorable setting for model evaluation. As the only publicly available tool that jointly supports outcome prediction and treatment-effect estimation in LVO stroke using both clinical and imaging features, MR PREDICTS serves as the benchmark for (model-assisted) prognostication relying on manual inputs.

To compare the results against a fully automated approach, we utilized a previously developed multimodal DL model [8]. This end-to-end approach directly incorporates initial raw CT angiography (CTA) images using transformer-based models [27] with pre-treatment clinical variables within an interpretable ordinal regression framework. Similar to MR PREDICTS, the model generates probabilistic predictions for ordinal mRS and supports individualized treatment-effect estimation without manual feature extraction, therefore representing a complementary, end-to-end approach to feature-based prognostication. Interpretability of clinical features is preserved through interpretable parameter estimates in terms of odds ratios. The model was trained on 449 patients from the MR CLEAN trial using five-fold cross-validation; a detailed introduction into the framework and its applications can be found elsewhere [28, 20].

### Neurologist prediction experiment

To assess neurologists’ ability to predict functional outcome and benchmark model performance, 40 cases were selected. Propensity score matching was used to balance patients treated with EVT and no-EVT. From the matched cohort, 20 pairs were randomly selected. Missing clinical variables were imputed using missForest [29].

Six neurologists from the University Hospital Zurich, with clinical experience in stroke care ranging from 4 to 20 years (median 10.5 years), participated as raters. All raters were blinded to patient outcomes and to each other’s predictions. Each rater was provided with the imputed clinical data (Table 1) and corresponding non-contrast CT and CTA DICOM images, which could be reviewed using a DICOM viewer of their choice. Neurologists were instructed to provide a full ordinal mRS prediction (0–6) for each patient. Outcome prediction was performed in two sequential rounds. In the first round, mRS predictions were based solely on clinical and imaging data. In the second round, raters repeated the prediction with access to MR PREDICTS and reported imaging-derived features as well as predicted treatment benefit for each case to ensure active use of the tool. Patient identifiers and case order were randomized independently between rounds to minimize recall bias.

**Table 1.** Descriptive statistics of the comparison cohort. The table summarizes frequencies (percentages) and mean (standard deviation) of outcome measures, imaging parameters and provided baseline data of the experimental test cohort.

|  | No-EVT (n = 19) | EVT (n = 21) |
| --- | --- | --- |
| **Outcome measures** |  |  |
| Favorable mRS 90d | 3 (15.8) | 7 (33.3) |
| mRS 90d |  |  |
| 0 | 0 (0.0) | 1 (4.8) |
| 1 | 1 (5.3) | 3 (14.3) |
| 2 | 2 (10.5) | 3 (14.3) |
| 3 | 2 (10.5) | 6 (28.6) |
| 4 | 7 (36.8) | 4 (19.0) |
| 5 | 5 (26.3) | 0 (0.0) |
| 6 | 2 (10.5) | 4 (19.0) |
| NIHSS 24h | 14.9 (5.8) | 11.2 (7.4) |
| **Imaging parameters** |  |  |
| Location occlusion |  |  |
| ICA-T | 10 (52.6) | 8 (38.1) |
| M1 | 9 (47.4) | 12 (57.1) |
| M2 | 0 (0.0) | 1 (4.8) |
| Collat. score |  |  |
| 0 | 1 (5.3) | 0 (0.0) |
| 1 | 6 (31.6) | 8 (38.1) |
| 2 | 5 (26.3) | 10 (47.6) |
| 3 | 7 (36.8) | 3 (14.3) |
| ASPECT | 8.5 (1.9) | 8.4 (1.2) |
| **Baseline data provided to raters** |  |  |
| IVT = yes | 16 (84.2) | 19 (90.5) |
| Age | 65.5 (13.2) | 60.4 (14.9) |
| Sex = male | 10 (52.6) | 11 (52.4) |
| NIHSS on adm. | 15.8 (6.0) | 17.2 (4.7) |
| Pre-stroke mRS = unfav | 2 (10.5) | 0 (0.0) |
| Time onset door (min) | 100.3 (90.0) | 101.2 (83.4) |
| Time onset IAT (min) | 250.5 (47.3) | 252.5 (76.5) |
| Time onset IVT (min) | 94.6 (55.7) | 94.3 (34.6) |
| Previous stroke = yes | 1 (5.3) | 1 (4.8) |
| Atrial fibrillation = yes | 5 (26.3) | 7 (33.3) |
| Diabetes = yes | 2 (10.5) | 2 (9.5) |
| Hypertension = yes | 8 (42.1) | 8 (38.1) |
| Systolic BP | 158.1 (27.8) | 148.4 (30.8) |
| Diastolic BP | 86.4 (19.4) | 83.1 (12.3) |
| Glucose | 7.6 (2.7) | 9.6 (10.3) |
| Creatinine | 79.2 (28.8) | 84.8 (21.9) |
| INR | 1.1 (0.3) | 1.1 (0.2) |

### Evaluation procedure

The primary outcome was functional disability measured by the ordinal mRS (0–6). For clinical interpretability, mRS was additionally dichotomized into favorable (mRS 0–2) and unfavorable (mRS 3–6) outcomes. Therefore, the model predicted probabilities of mRS classes 0–2 were summed up. In case of the validated MR PREDICTS, the prediction was classified as favorable when the probability exceeded a probability threshold of 0.5. For the DL model, this threshold was learned using the training and validation data of the respective cross-validation fold using Youden’s index.

Descriptive statistics were used to summarize the comparison cohort (n=40). Frequencies and percentages respectively means and standard deviations were used to describe outcome measures, baseline data and imaging parameters for patients with favorable and unfavorable outcome.

Neurologists’ predictions were then compared to those of the models in the comparison subset. To provide descriptive context for model performance, we additionally evaluated the models in complete cases of the MR CLEAN trial available to both models (n=404 patients). The DL model was always evaluated using the test data.

Model-predicted probabilities for binary mRS were assessed in terms of discrimination and calibration using the area under the receiver operating characteristics curve (AUC) and Brier score. Neurologists’ predicted classes of ordinal mRS were compared to those of the models using quadratic weighted kappa (QWK) and accuracy within a tolerance range of ±1 mRS point. While QWK measures agreement across the ordinal mRS, penalizing larger discrepancies more heavily, ordinal accuracy evaluates how often predicted mRS was within one point of the true mRS. Binary outcome predictions were evaluated using accuracy, sensitivity (correctly predicted favorable outcomes), and specificity (correctly predicted unfavorable outcomes). Agreement within individual raters across prediction rounds was assessed using QWK for ordinal mRS and Cohen’s kappa for binary mRS. Agreement between raters within each prediction round was summarized using Fleiss’ kappa.

### Error and input estimation analysis

Prediction error, defined as the difference between predicted and observed mRS, was evaluated both across all neurologists combined as well as at the individual rater level. Factors associated with systematic biases were explored using linear mixed-effects models with prediction error as outcome, pre-treatment clinical variables as predictors and a random intercept for each rater. Neurologists’ ability to estimate imaging-derived features required for MR PREDICTS was evaluated using accuracy for occlusion location and collateral score, and root mean squared error (RMSE) for ASPECTS.

### Uncertainty quantification and Code availability

Uncertainty in performance metrics was quantified using a clustered bootstrapping approach with 1000 resamples. Sampling was performed with replacement at the patient level rather than the individual prediction level, ensuring that all associated rater assessments for a given patient were drawn together to account for the study design. All statistical analyses were performed using R version 4.5.2 and the code is publicly available on Github via this link https://github.com/liherz/prediction_challenge_mrclean/tree/main.

## Results

### Study population

In the comparison subset, baseline characteristics were well balanced across groups, including age, sex, vascular risk factors, pre-stroke mRS, admission NIHSS, imaging characteristics, and laboratory values (Table 1). Most patients in the treatment and control group received IVT. Occlusions were primarily located in the ICA terminus (ICA-T) or M1 segment. A favorable outcome was achieved in 33.3% of EVT treated patients and in 15.8% of the control group, representing a comparable treatment effect (Odds Ratio 2.67) as observed in the full MR CLEAN trial cohort.

### Model performance in the larger cohort (n=404 patients)

MR PREDICTS and the DL model showed comparable performance for both ordinal mRS and binary outcome prediction (Table 2).

**Table 2:** Summary of ordinal and binary prediction performances. Prediction performances of models and neurologists for the different cohorts of size n. Ordinal mRS: quadratic weighted kappa (QWK), (±1 mRS point) accuracy. Binary mRS: area under the receiver operating characteristics curve (AUC), brier score, accuracy (Acc), sensitivity (Sens) and specificity (Spec).

|  | **MR PREDICTS** |  | **DL MODEL** |  | **Neurologists** |  |
| --- | --- | --- | --- | --- | --- | --- |
|  | **Complete cases**<br>**(n=404)** | **True imaging features**<br>**(n=40)** | **Complete cases**<br>**(n=404)** | **True imaging data**<br>**(n=40)** | **Without MR PRED**<br>**(n=40)** | **With MR PRED**<br>**(n=40)** |
| QWK | 0.48<br>[0.41, 0.54] | 0.51<br>[0.24, 0.70] | 0.41<br>[0.33, 0.50] | 0.49<br>[0.25, 0.67] | 0.27<br>[0.10, 0.42] | 0.41<br>[0.22, 0.55] |
| Acc<br>($\pm 1$) | 55.69<br>[50.74, 60.40] | 65.00<br>[50.00, 77.50] | 62.38<br>[57.67, 67.33] | 63.16<br>[47.37, 78.95] | 58.75<br>[48.33, 68.34] | 57.50<br>[48.33, 66.25] |
| AUC | 0.78<br>[0.73, 0.83] | 0.77<br>[0.59, 0.92] | 0.74<br>[0.69, 0.80] | 0.77<br>[0.42, 0.91] | - | - |
| Brier | 0.20<br>[0.18, 0.22] | 0.17<br>[0.12, 0.22] | 0.17<br>[0.15, 0.19] | 0.17<br>[0.11, 0.24] | - | - |
| Acc | 66.83<br>[62.13, 71.53] | 67.50<br>[52.50, 82.50] | 71.29<br>[67.07, 75.74] | 63.16<br>[47.37, 78.95] | 64.17<br>[55.42, 72.92] | 68.75<br>[58.33, 78.34] |
| Sens | 80.19<br>[72.28, 87.50] | 60.00<br>[27.22, 88.89] | 66.98<br>[57.98, 76.24] | 70.00<br>[40.00, 100.00] | 61.67<br>[45.45, 76.68] | 66.67<br>[45.45, 85.71] |
| Spec | 62.08<br>[56.78, 67.48] | 70.00<br>[53.33, 86.21] | 72.82<br>[67.93, 77.74] | 60.71<br>[42.29, 78.27] | 65.00<br>[54.99, 75.01] | 69.44<br>[57.64, 80.25] |

Ordinal mRS predictions were closer to the true mRS classes for MR PREDICTS (QWK 0.48 [0.41 to 0.54]) than for the DL model (QWK 0.41 [0.33 to 0.50]), while DL model predictions were more frequently within +/- one point of the true mRS class (ordinal accuracy 62.38% [57.67% to 67.33%] vs. 55.69% [50.74% to 60.40%]). For binary mRS prediction, the DL model achieved a higher accuracy of 71.29% [67.07% to 75.74%] vs. 66.83% [62.13% to 71.53%] for MR PREDICTS.

Regarding discriminative performance, i.e. the models’ ability to distinguish patients with favorable vs. unfavorable outcome using the predicted probabilities, MR PREDICTS showed a slightly higher AUC of 0.78 [0.73 to 0.83] (vs. 0.74 [0.69 to 0.80]). However, the DL model demonstrated superior calibration (Brier score 0.17 vs. 0.20), indicating more reliable probability estimates.

### Model performance in the comparison cohort (n=40 patients)

In the comparison subset, the standalone performances of both models remained stable (Table 2). Ordinal agreement was nearly identical (QWK MR PREDICTS 0.51 [0.24 to 0.70]; QWK DL model 0.49 [0.25 to 0.67]), as was ordinal accuracy (MR PREDICTS 65.00% [50.00% to 77.50%] vs. DL model 63.16% [47.37% to 78.95%]). For binary mRS, MR PREDICTS achieved an accuracy of 67.50% [52.50% to 82.50%] compared to 63.16% [47.37% to 78.95%] for the DL model. MR PREDICTS achieved a higher sensitivity, the DL model a higher specificity; yet, values were largely balanced. The overall discriminative performance and calibration for binary mRS probabilities was similar, with an AUC of 0.77 and a Brier score of 0.17 for both.

### Neurologists’ performance

Without prognostic support, the six neurologists’ ordinal mRS predictions showed high variability and significantly lower agreement with observed outcomes compared to the standalone models (QWK 0.27 [0.10 to 0.42]; ordinal accuracy 58.75% [48.33% to 68.34%]; Table 2, Figure 2). This variability was observed across all mRS classes among all raters combined (Figure S1) and at the individual rater level (Figure S2 and S3), resulting in poor interrater consensus (Fleiss’ κ=0.11). In contrast, binary accuracy for predicting a favorable vs. unfavorable outcome was closer to model performance (64.17% [55.42% to 72.92%]), showing fair interrater agreement (Fleiss’ κ=0.25).

**Figure 2:**
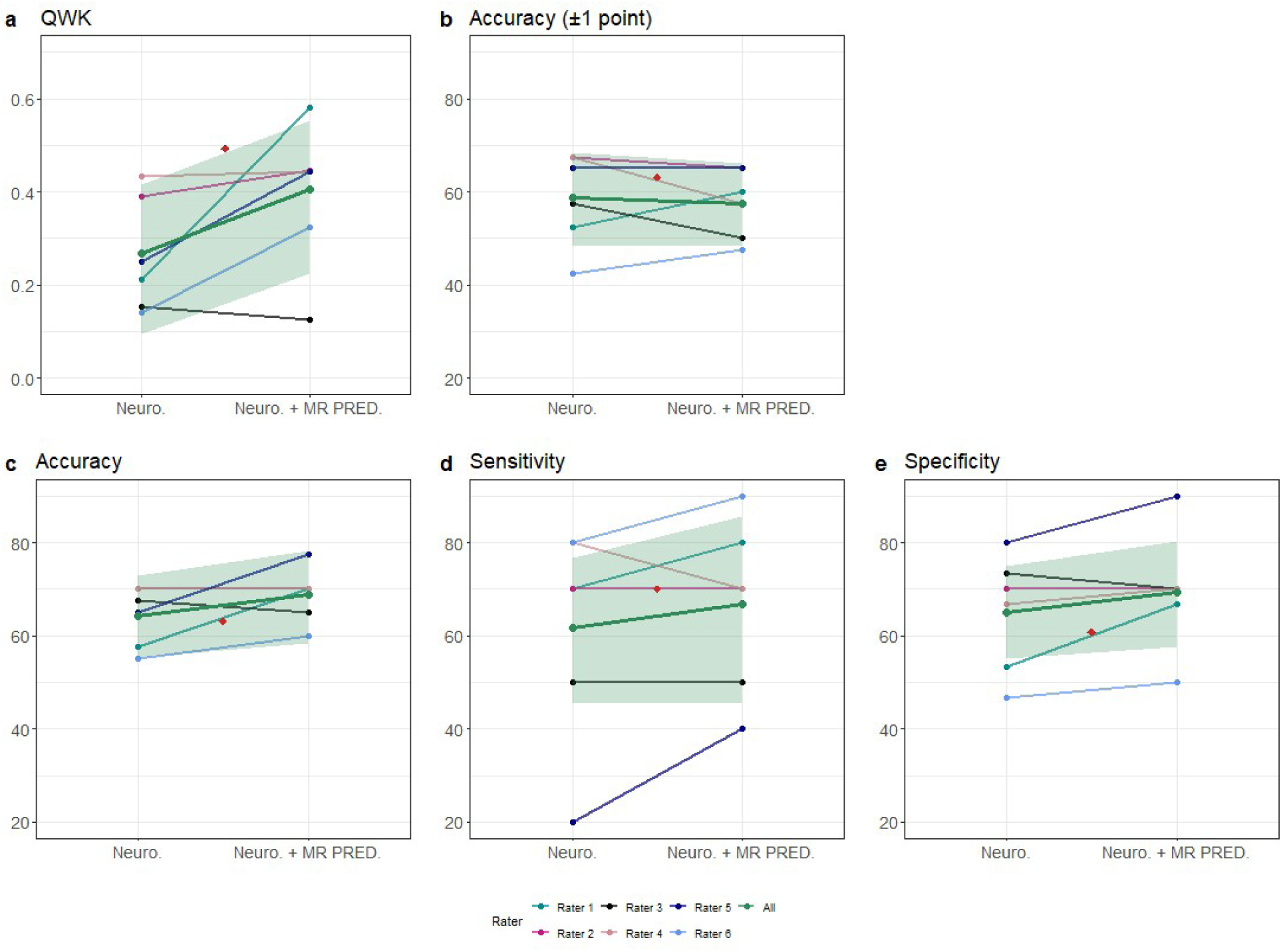
Prediction performance of neurologists for ordinal and binary mRS. The figure shows the mean prediction performance with 95% CI alongside individual prognostications of neurologists in terms of a) quadratic weighted kappa (QWK), and b) accuracy (± 1 mRS point) for ordinal mRS as well as c) accuracy, d) sensitivity, and e) specificity for binary mRS. Performance is shown for predictions without (Neuro.) and with (Neuro. + MR PRED) access to MR PREDICTS. The performance of the DL model is integrated with a red dot.

Access to MR PREDICTS modestly improved average prediction performance, which, however, remained below that of the standalone models in the ordinal case. While ordinal mRS predictions were closer to the true ones (QWK 0.41 [0.22 to 0.55]), ordinal accuracy slightly decreased to 57.50% [48.33% to 66.25%], with interrater agreement remaining low (Fleiss’ κ=0.11). Notably, however, access to the tool improved the clinicians binary prediction accuracy to 68.75% [58.33% to 78.34%] and substantially harmonized their predictions as indicated by a higher Fleiss’ κ of 0.40. Within-rater agreement between prediction rounds was moderate to substantial for both ordinal (QWK 0.49 to 0.71) and binary outcomes (Cohen’s κ 0.39 to 0.59).

### Systematic biases and limitations in human prediction

Neurologists’ systematically overestimated functional outcomes, predicting lower (more favorable) mRS classes than were actually observed. This optimism bias was present across all raters and at the individual rater level, both with and without access to MR PREDICTS. The models showed symmetric error distributions (Figure 3, Figure S4 and S5). Estimating the imaging features required for the MR PREDICTS proved highly challenging for the clinicians. On average, occlusion location was correctly identified in 65.8% [57.1% to 74.6%] of cases; collateral score accuracy was lower with 44.6% [36.7% to 52.5%]. ASPECTS predictions deviated by 3.4 points [3.1 to 3.6] from the confirmed estimates, accompanied by substantial interrater variability (Table S1).

**Figure 3:**
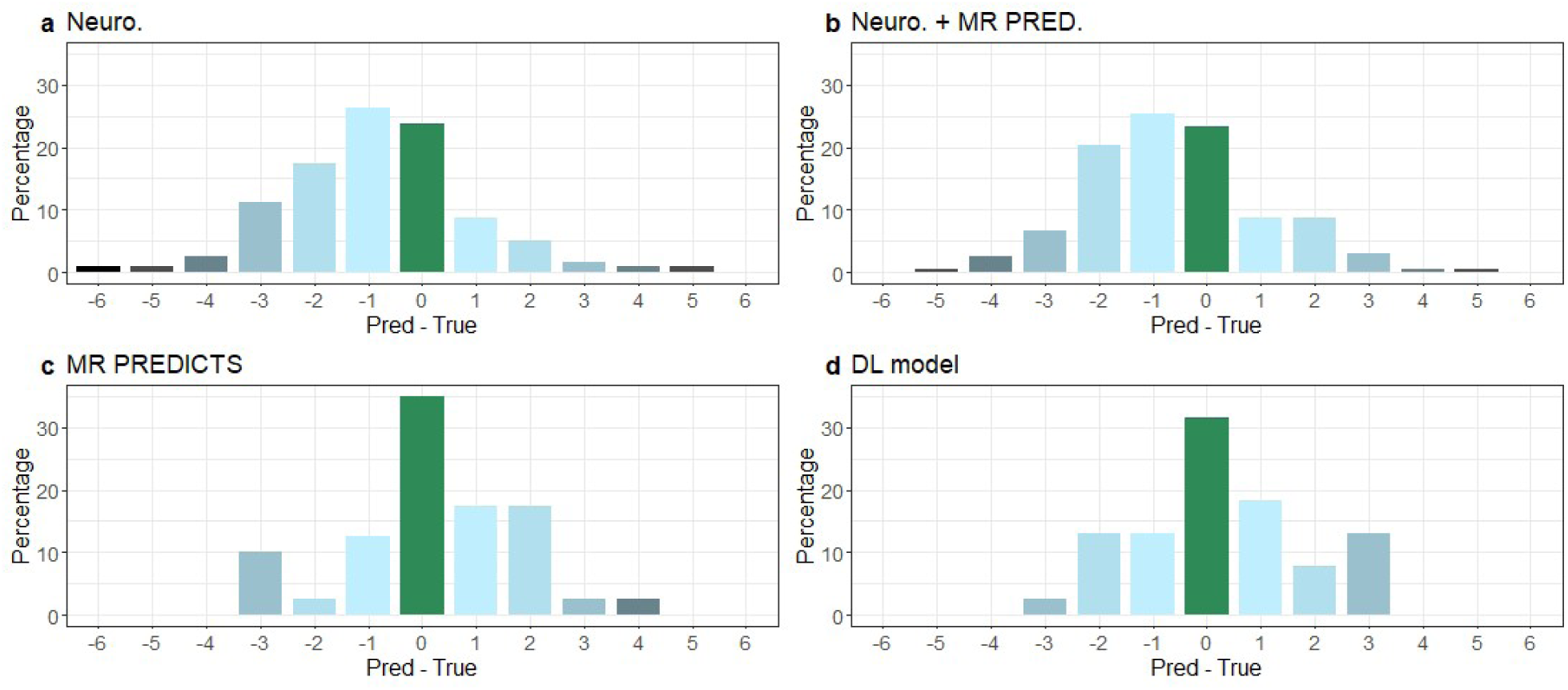
Prediction errors of neurologists and models for ordinal mRS. The figure shows prediction errors of neurologists (Neuro.) by summarizing the difference between predicted and true mRS outcomes a) without and b) with access to MR PREDICTS as well as the errors of c) MR PREDICTS and d) DL model in the comparison subset. Negative values indicate optimism, positive values pessimism.

### Factors associated with prediction error

Linear mixed-effects models with random intercepts for the raters (Table S2) identified specific patient characteristics associated with prediction errors. Clinicians tended towards pessimism in male patients (effect size 0.90 [0.45 to 1.35]) and those with hypertension (0.77 [0.25 to 1.29]) by estimating higher mRS scores than observed. The initiation of acute therapies was associated with overly optimistic predictions for both EVT (−0.23 [−0.65 to 0.19]) and IVT (−0.35 [−1.05 to 0.35]) treated patients. Other variables, including age, NIHSS, systolic blood pressure, diabetes, glucose, and onset-to-door time, showed no consistent association with human error. These patterns remained similar when clinicians had access to MR PREDICTS (s. also Table S3 and S4).

## Discussion

In this study, we systematically compared neurologists, features-based models and DL for outcome prediction in LVO stroke. For ordinal mRS outcomes, models significantly outperformed neurologists while performance for binary mRS was comparable. Model-assistance yielded only moderate improvements in neurologists’ predictions, indicating a prognostic gap where predictive superiority of a model does not automatically translate into enhanced human performance.

MR PREDICTS and the DL model demonstrated moderate predictive performance in the complete-case and the comparison subset, with AUC values around 0.77. This is consistent with previously reported AUC ranges for multimodal prognostic models in ischemic stroke [21, 30, 22, 31]. Importantly, when interpreting these findings, it must be noted that MR PREDICTS was evaluated on its own training data, potentially inflating its performance due to overfitting. In contrast, the end-to-end DL model achieved a comparable AUC despite being strictly evaluated on unseen test data, underlining its robustness. Crucially, neurologists’ unaided predictions showed significantly lower ordinal accuracy and considerable interrater variability, with a systematic tendency toward overestimating favorable functional outcomes (optimism bias), particularly in patients treated with IVT and EVT. This suggests that clinical intuition may be biased by treatment-effect expectations, leading clinicians to overestimate the likelihood of recovery. While MR PREDICTS partially aligned neurologists’ predictions, overall performance remained below that of the standalone models. This suggests that model anchoring or a lack of trust in model output may hinder effective human-model collaboration.

Despite the availability of structured and imputed trial data, reflecting an ideal scenario compared with real-world conditions, neurologists’ ordinal and binary performance was highly variable. This highlights the difficulty of assessing pre-treatment clinical and CT imaging data. In addition, the moderate performance of both models and humans likely reflects a biological upper bound on achievable performance for mRS prediction: events occurring after the acute phase such as recurrent stroke or medical complications cannot be captured by baseline data. Future research should therefore explore treatment-conditional outcome measures that better reflect individualized recovery trajectories.

A primary driver of human bias was imaging assessment. Consistent with previous observations [11], neurologists struggled to accurately interpret imaging data, here regarding the estimation of manual imaging features required by feature-based models. This error-prone manual extraction limits the clinical utility of such models. In contrast, end-to-end DL models directly integrate raw imaging data with clinical variables, eliminating the subjectivity of human imaging interpretation and feature estimation while achieving comparable or superior predictive performance.

Several limitations should be acknowledged. First, the neurologist prediction experiment involved a relatively small comparison cohort to ensure feasibility. Additionally, the DL approach requires further external validation to ensure generalizability to current LVO stroke patients. As notes above, the MR CLEAN trial data provides a favorable setup for MR PREDICTS, limiting the direct comparison with the DL model that was evaluated on the test data. The cohort data (2010-2014) predates recent interventional procedures and devices. However, the persistence of unfavorable outcomes in approximately 50% of patients suggests that the fundamental challenge of functional outcome prognosis remains highly relevant [33]. Finally, while our study design provided a controlled comparison aiming to mimic real world prognosis, it cannot fully replicate the acute time-pressure and neurologists’ intuition derived from direct patient contact in real world care. Additional prospective evaluations are essential to assess the real-world impact of outcome prediction models on clinical decision-making and integration.

In conclusion, the multimodal models significantly outperformed neurologists in predicting the full ordinal mRS. While the modest improvement in model-assisted performance underscores that clinical utility depends on successful human-machine interaction, our findings demonstrate that end-to-end DL approaches successfully overcome the critical bottleneck of manual imaging-feature variability. By providing rapid and objective prognostic baselines directly from raw imaging data, advanced DL models hold immense potential to serve as a reliable, second opinion. Instead of acting as gatekeepers to withhold treatment, they should be considered as supporting tools for clinicians to optimize individualized prognostication for improved decision-making and family counseling in acute stroke care.

## Data Availability

The MR CLEAN trial cohort is available from the MR CLEAN consortium but restrictions apply to the availability of the data, which were used under license for the current study, and so are not publicly available. Data are however available from the authors upon reasonable request and with permission of the MR CLEAN consortium.

## Supplemental Material

Table S1-S4 Figure S1-S5

## Acknowledgments

None.

